# Solitary Lung Cavities on CT Imaging: Differentiating Malignant and Non-Malignant Diseases

**DOI:** 10.1101/2024.06.09.24308668

**Authors:** Zeliha Cosgun

## Abstract

**Background:** This study aims to explore how radiological findings contribute to distinguishing between benign and malignant diseases in patients with solitary cavitary lesions detected on CT.

**Methods:** We retrospectively assessed lesion size, cavity wall thickness, and additional parenchymal findings in diagnosing benign or malignant disease in these patients. Our study investigated the incidence and etiology of solitary pulmonary cavities. CT scans were reviewed by a single radiologist with expertise in thoracic radiology. The study was conducted using two 64-multidetector CT systems, and measurements of lesion size and cavity wall thickness were recorded on axial images. Consolidation and centrilobular nodules were assessed based on predefined criteria. Receiver operating characteristic curves were generated to determine optimal cut-off points for distinguishing between malignant and non-malignant lesions based on cavity wall thickness.

**Results:** Non-malignant lesions accounted for 47.9% of cases, with active pulmonary tuberculosis being the most common diagnosis. In the malignant group, primary lung cancer predominated, squamous cell carcinoma being the most prevalent subtypes. Significant differences were noted between malignant and non-malignant cases regarding average maximum wall thickness and lesion diameter. Presence of perilesional consolidation or centrilobular nodules favored non-malignant diagnoses. Maximum wall thickness thresholds of 7.2 mm or 23 mm were most accurate in suggesting non-malignant and malignant etiologies, respectively.

**Conclusions:** In conclusion, CT findings revealed significant distinctions between malignant and non-malignant solitary lung cavities; benign lesions generally exhibited smaller and thinner cavity walls, with accompanying perilesional parenchymal findings observed in benign lesions of infectious origin but not in malignant lesions.

## Introduction

A lung cavity is characterized as a gas-containing lesion within the lung, enclosed by a wall of variable thickness. An excavation is defined as the manifestation of a cavity, which may be situated within an opacity such as consolidation, mass, or nodule. Various mechanisms can lead to the formation of cavities, with the most common being tissue loss due to necrosis within a mass or nodule, which can be neoplastic, infectious, or ischemic in natüre [1].

Cavitated lung lesions are commonly observed on imaging studies, with malignancies and infections representing the predominant origins. These lesions stem from various causes, leading to diverse etiologies and diagnostic challenges due to shared imaging features.

The spectrum of potential causes for cavitary lung lesions is extensive and encompasses infectious diseases like tuberculosis, fungal infections, and parasitic infections, as well as noninfectious conditions such as malignant and rheumatic lesions [2-4].

Conventional chest radiography and computed tomography (CT) are the primary imaging modalities utilized for pulmonary disease assessment., CT emerges as the most dependable and sensitive technique for cavitary lung lesion evaluation.

In solitary cavity lesions, multiple CT findings such as cavity diameter, cavity wall thickness, and the presence of parenchymal involvement surrounding the lesion aid in diagnosis. A recent study found that a cavity wall thickness of less than 7 mm is highly specific for benign disease, while thickness exceeding 24 mm is highly specific for malignant disease [5]. Additionally, the absence of perilesional parenchymal findings often supports malignancy, while their presence often suggests benignity [3].

The acute onset of symptoms can sometimes assist in distinguishing between malignant and non-malignant diseases. For example, a benign infection may cause hemoptysis when it affects a nearby vessel. Benign diseases can also lead to fatigue and weight loss similar to malignancies. The acute onset of fever generally helps differentiate benign disorders from malignancies, but lung cancer can manifest with a secondary superinfection to the tumor [6].

Therefore, correlating clinical and laboratory findings with radiological findings is essential in distinguishing between benign and malignant lesions. Preliminary diagnoses and differential diagnoses established based on radiological findings could be crucial for facilitating early diagnosis. Hence, the aim of our study is to evaluate diagnostic CT findings in benign and malignant cavitary lung lesions.

## Methods

In our cross sectional study, we retrospectively evaluated 186 patients with cavitary lung disease described on CT scans between 2015 and 2021. Among them, 105 patients with multiple cavitary lung lesions and no clinical-laboratory or pathological diagnosis were excluded. And 2 case of fibrocavitary tuberculosis in the upper lobe, 6 cases of lobar necrotizing pneumonia were excluded (figüre 1). Only 73 patients with solitary cavitary lesions and a confirmed diagnosis through clinical, laboratory, pathological findings, or follow-up imaging were included in our study. The images of these patients were reevaluated by a single radiologist with 9 years of experience in thoracic radiology and a total of 17 years of radiological experience, using the Picture Archiving and Communication System (PACS). This study was approved by our hospital’s ethics committee (2021-233).

**Figure 1:**
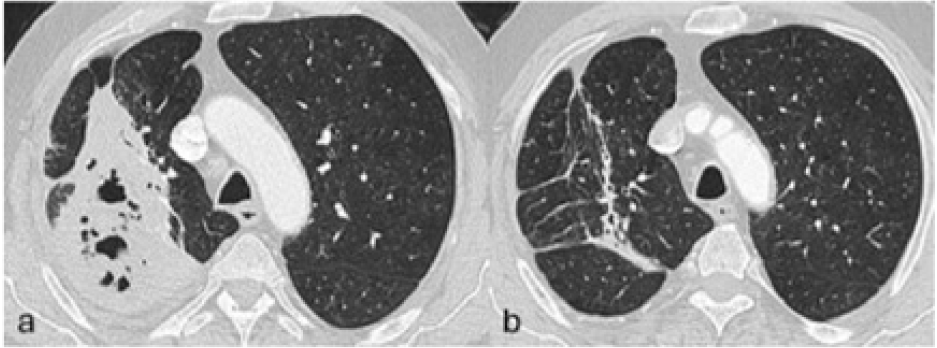
Thoracic CT, axial images in parenchymal window, Consolidation areas with air bronchograms and accompanying cavities, located in the posterior aspect of the right upper lobe and the superior segment of the right lower lobe, consistent with necrotizing pneumonia(a). Follow-up imaging after one year: Regression of pneumonic infiltrates with residual changes and volume loss in the same localization, indicative of sequelae (b).

Examinations were conducted using two 64-multidetector CT systems (General Electric Revolution EVO, 64 slices). CT scans were performed without ECG gating, with the following scan parameters: 0.6 mm collimation, 1.5 mm slice thickness, 1.4 mm increment, 100 kV, 135 mAs, a pitch of 0.9, and a gantry rotation time of 0.33 s. Intravenous administration of 100 cc nonionic contrast material was done using an automatic injector at a flow rate of 4 mL/s via a vein in the arm.

Each examination involved recording the lesion’s size (maximum diameter) and the maximum wall thickness of the cavity, both measured on axial images, along with any associated findings, regardless of their location or extent. Images were volumetrically acquired and assessed using high-resolution and soft kernels on both pulmonary and mediastinal windows, respectively. Measurements were specifically conducted in the axial plane on the pulmonary window to enhance result reproducibility and employ a readily applicable technique.

Consolidation was defined as a uniform increase in lung attenuation that obscured the margins of adjacent vessels and airway walls. Centrilobular nodules were identified as nodular opacities situated at the center of a normal secondary pulmonary lobule, following the guidelines set forth by the Nomenclature Committee of the Fleischner Society.

The Statistical Package for the Social Sciences (SPSS) software (version 15, SPSS Inc., Chicago, IL, US) for Windows was used for statistical analysis. Patients were categorized into two groups based on their final diagnosis: malignant and non-malignant etiologies of solitary pulmonary cavities. Group variables were assessed using independent-sample Student’s t-tests for numerical data with a normal distribution, and Chi-square tests for categorical data or proportions. All tests were two-tailed, with significance set at p<0.05. Following initial analysis, receiver operating characteristic (ROC) curves were generated to identify optimal cut-off points for distinguishing between malignant and non-malignant lesions based on maximum wall thickness of the cavities.

## Results

Upon reviewing CT images and medical records, 73 patients with solitary pulmonary cavities (35 benign, 38 malignant) were identified. The average age for benign lesions was 58±14 years, whereas for malignant lesions it was 64±9 years. Among the total of 73 patients, 15 (13.7%) were female. Of those with benign lesions, 10 were female and 25 were male, while among those with malignant lesions, 5 were female and 33 were male.

Non-malignant lesions accounted for 35 cases (47.9%), while 38 cases (52.1%) were malignant. Among the non-malignant diagnoses, mycobacterial infection sequelae comprised 5 cases (14.3%) (figüre 2), active pulmonary tuberculosis 11 cases (31.4%) (figüre 3), fungal infections 4 cases (11.4%) (figüre 4a and 4b), non-fungal infections 12 cases (34.3%) (figüre 5a-b,6,7,8) and rheumatoid nodules 3 cases (8.6%) (figüre 9).

**Figure 2:**
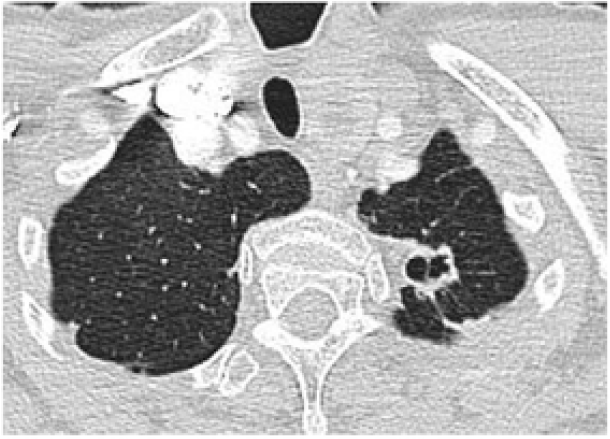
In a patient with a history of previous tuberculosis (TB), a cavitary lesion with a stable thin wall in the apicoposterior segment of the left lung was interpreted as consistent with a sequel TB cavity.

**Figure 3:**
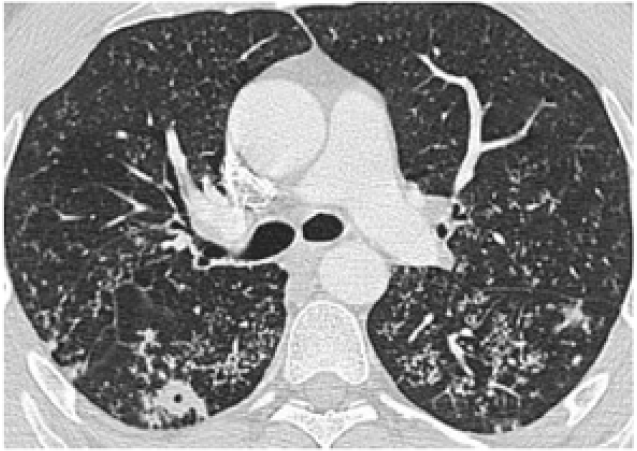
In a patient diagnosed with active tuberculosis (TB), a cavitary lesion in the superior segment of the right lower lobe of the lung and widespread tree-in-bud opacities in the bilateral lung parenchyma, consistent with endobronchial spread, were observed.

**Figure 4:**
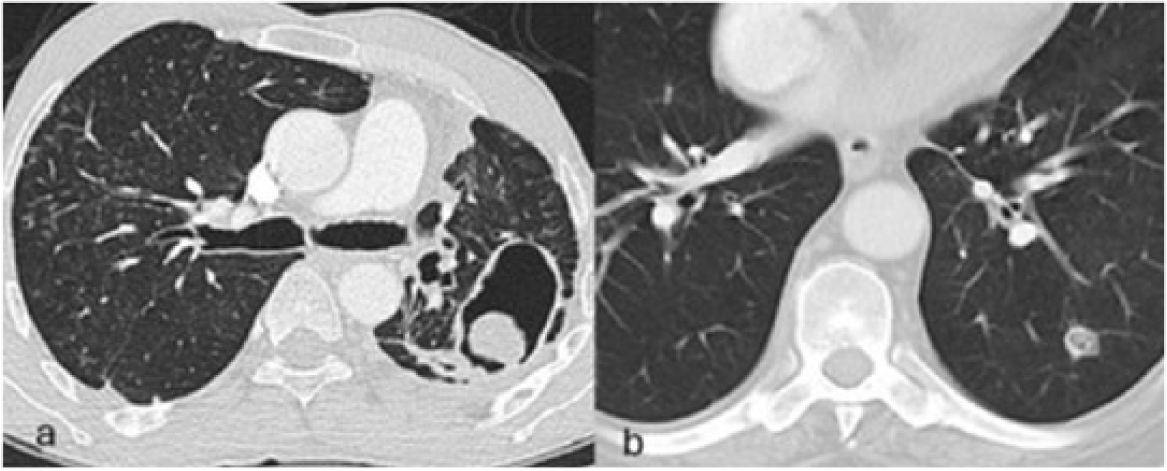
Fungal infections, In a patient diagnosed with Aspergillus fumigatus, a fungus ball within the cavity was observed in the left lung following left upper lobectomy(a). In a patient diagnosed with Candida albicans, a cavitary lesion containing a solid component was observed in the left lower lobe of the lung(b).

**Figure 5:**
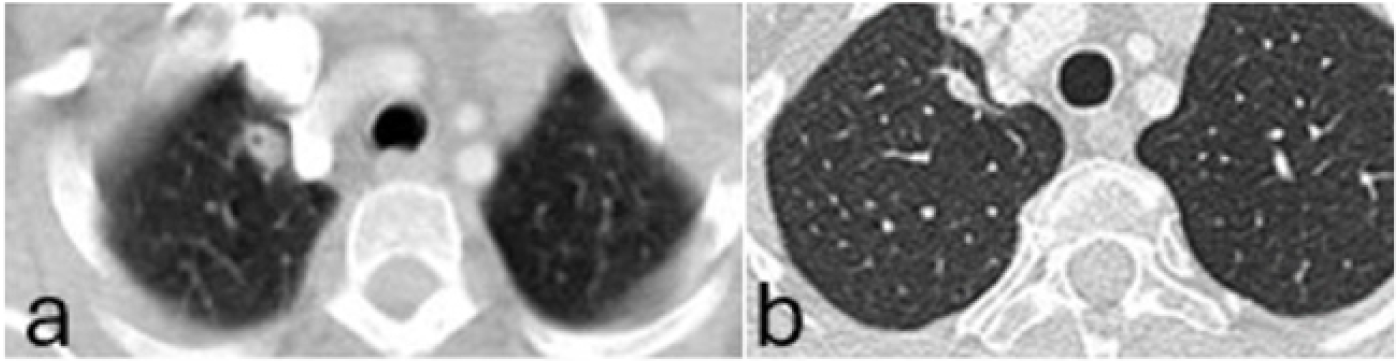
A cavitary lesion observed in the right lung apex (a), patient followed up with a diagnosis of hydatid cyst, demonstrates regression (b) after hydatid cyst treatment.

**Figure 6:**
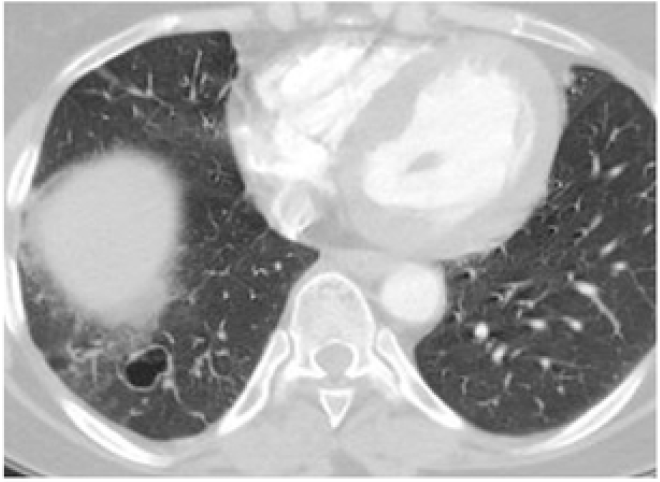
In a patient diagnosed with Streptococcus pneumoniae, a cavitary lesion in the posterior basal segment of the right lower lobe of the lung accompanied by perilesional ground-glass opacities.

**Figure 7:**
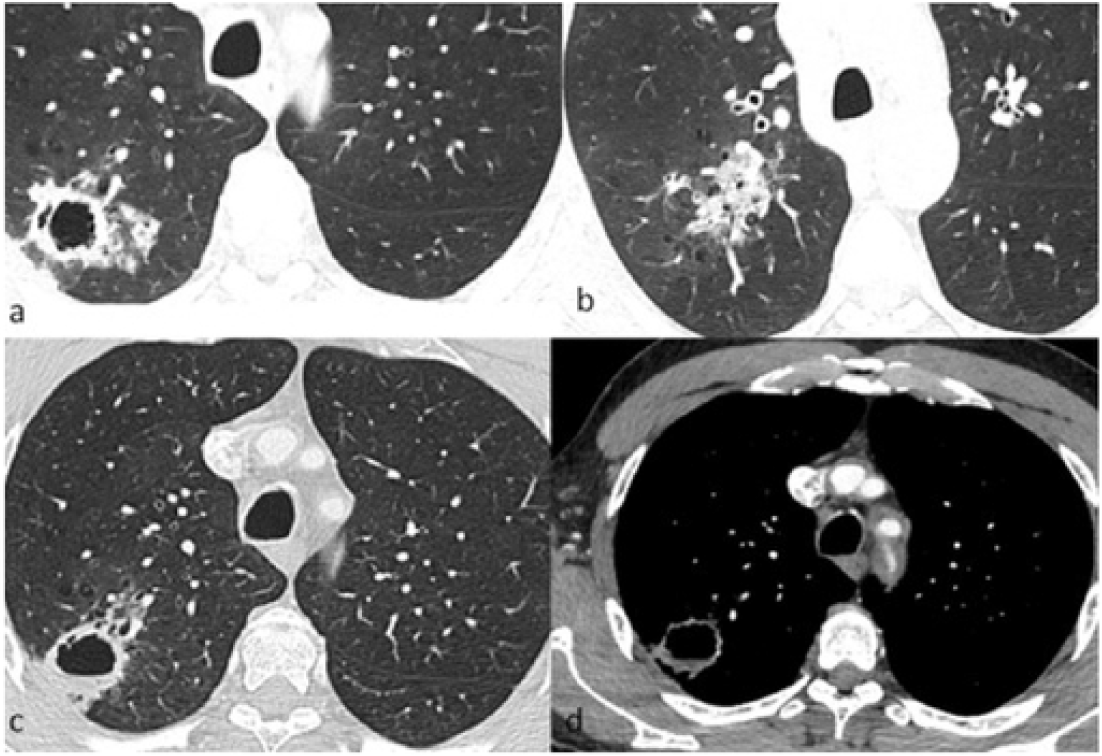
In a patient diagnosed with Nocardia, a cavitary lesion accompanied by perilesional consolidation was observed in the right lower lobe of the lung.

**Figure 8:**
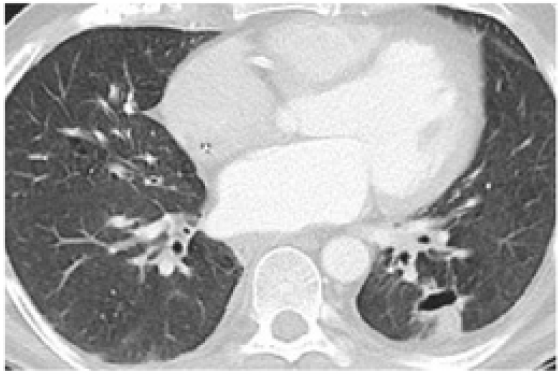
In a patient diagnosed with nontuberculous mycobacteria, a thin-walled cavitary lesion is observed in the left lower lobe of the lung.

**Figure 9:**
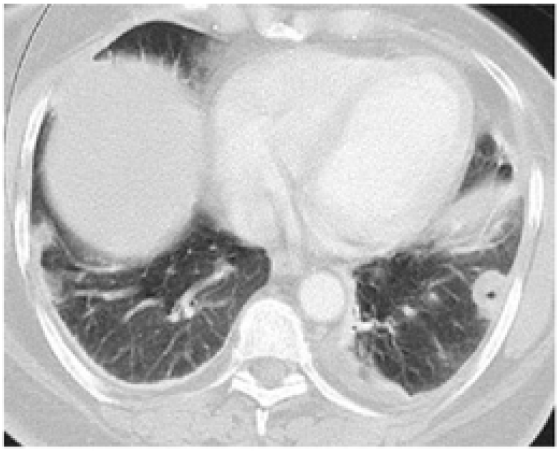
A cavitary lesion consistent with rheumatoid nodule, pathologically diagnosed in the left lower lobe of the lung

Out of the 12 patients diagnosed with non-fungal infections, 2 were under follow-up with a diagnosis of hydatid cyst (figüre 5a-b), 1 had Streptococcus pneumoniae (figüre 6) as the pathogen, 2 had Nocardia (figüre 7), and 1 had a bacterium other than the Mycobacterium tuberculosis complex (figure8). In 6 patients, regression of the lesion on follow-up imaging led to the consideration of an infectious etiology for the lesion. Among the 4 patients diagnosed with fungal infections, Aspergillus fumigatus was detected in 2 cases (figüre 4a), and Candida albicans in 2 cases (figüre 4b).

In the malignant group, primary lung cancer was diagnosed in 32 patients, while 6 patients (15.8%) had metastases (figüre 10a). Among primary lung cancers, there were 9 cases of adenocarcinoma (23.7%) (figüre 10b), 23 cases of squamous cell carcinoma (60.5%) (figüre 10c). All malignant cases were histologically confirmed, whereas non-malignant cases were confirmed by clinical/radiological and laboratory findings, as well as follow-up imaging.

**Figure 10:**
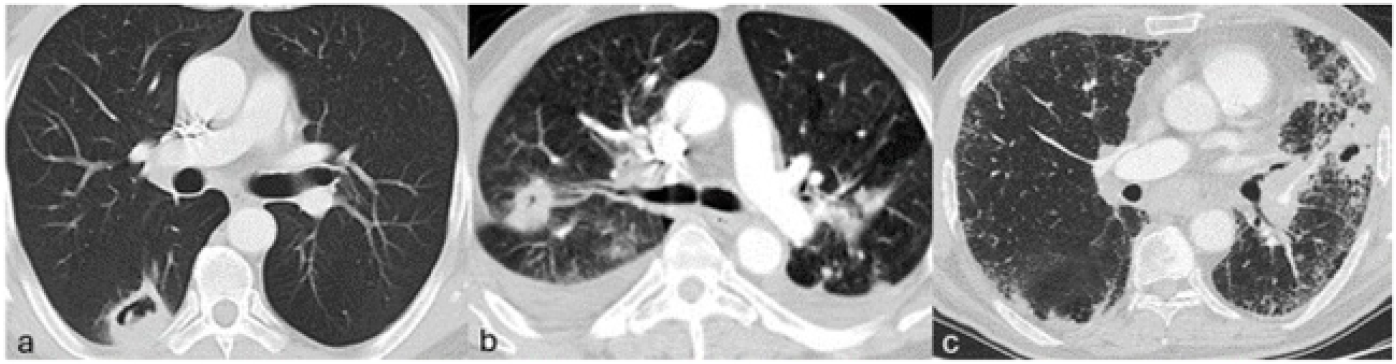
In a patient followed up with a diagnosis of bladder cancer, pathologically diagnosed with metastasis, a single cavitary lesion is observed in the superior segment of the right upper lobe of the lung (a). Primary lung adenocancer (b) and Squamous cell carcinoma in the background of UIP pattern (c).

While benign lesions are most commonly observed in the apical segment (12 patients 34.3 %) of the right upper lobe, malignant lesions are most frequently detected in the anterior-posterior segment of the right upper lobe (8 patients 21.1 %).

In 11 out of 35 patients with benign lesions, accompanying perilesional consolidation areas and centrilobular nodules were observed. While perilesional parenchymal findings were not detected in malignant lesions, accompanying satellite nodules were observed in 6 patients (p<0.01).

The maximum lesion diameter averages differed significantly between malignant and non-malignant groups. In the malignant group, the longest dimension was 47.7 mm (25-70), while in the benign group it was 30.9 mm (16-42) (p<0.01). The maximum wall thickness at the thickest point of the cavity was 20.6 mm (8.8-35) in malignant cases, whereas it was 6.8 mm (3.8-7) in benign cases (p<0.01).

Thresholds for maximum wall thickness associated with non-malignant and malignant lesions were identified: 7.2 mm was 84% specific and 77% sensitive for benignity, while 23 mm achieved a specificity of 97% and a sensitivity of 45% for malignant lesions (figüre 11).

**Figure 11:**
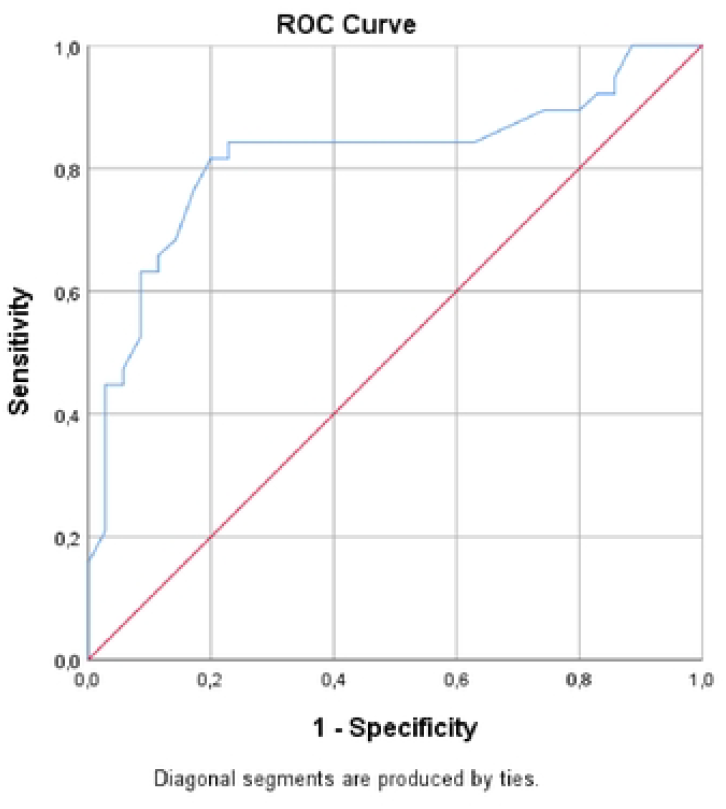
ROC analysis in maxmnum wall thickness associated with malignant cavities lesions

## Discussion

The most significant result of our study is that lesion size and cavity wall thickness are significantly higher in malignant lesions compared to benign lesions. The study identified cutoff values for maximum wall thickness associated with non-malignant and malignant lesions: 7.2 mm for benignity and 23 mm for malignancy. Similar to our study, many studies have emphasized the importance of imaging findings of cavitary lesions in contributing to diagnosis. Additionally, numerous studies in the literature have identified significant differences in cavity wall thickness between benign and malignant cavitary lesions, findings which are consistent with our study. According to Woodring et al. A wall thickness exceeding 15 mm indicated a likelihood of malignancy in X-ray scans [7,8], while Nin et al. Set the cutoff at greater than 24 mm for CT scans [3].

Our study findings, in line with existing literature, suggest that increased cavity wall thickness in pulmonary lesions correlates with malignancy, likely associated with the invasive behavior of tumor cells, tissue damage, and the resultant inflammatory response due to necrosis. Previous research has indicated that heightened cavity wall thickness in malignant lesions may stem from the aggressive nature of tumor cells, which infiltrate surrounding tissues, leading to localized tissue damage and subsequent inflammation. Furthermore, the observed inflammatory response may be attributed to the release of cytokines and other inflammatory mediators triggered by tumor cell necrosis and apoptosis. Additionally, emerging evidence suggests that the inflammatory milieu within the cavity may contribute to immune suppression, thereby facilitating tumor progression and metastasis. The association between increased cavity wall thickness and malignancy underscores the complex interplay between tumor biology and the host microenvironment, emphasizing the potential utility of cavity wall thickness as a diagnostic marker for malignancy in pulmonary lesions.

Another significant finding of our study, consistent with the literature, is the presence of perilesional centrilobular nodules and consolidations accompanying benign lesions [9-11].

In our study, the finding that perilesional consolidation observed in cavitary lesions supports benignity emerged as a significant observation. Consistent with the literature, perilesional consolidation is commonly associated with benign lesions, often attributed to infections, granulomatous diseases, and reactive changes. In this context, the presence of perilesional consolidation may decrease the likelihood of malignant lesions and support the benign nature of the lesion. However, it is crucial to note that perilesional consolidation alone is not sufficient for diagnosis, and clinical findings, imaging features, and histopathological examination when necessary should all be considered. This finding could serve as an additional evaluation criterion in the assessment of cavitary lesions in clinical practice, contributing to achieving more accurate diagnoses in the diagnostic process.

It’s important to acknowledge certain limitations of this study. Firstly, due to its retrospective design, the authors were unable to rectify occasional inaccuracies in the databases. Secondly, the relatively small sample size precluded the possibility of conducting an analysis of covariance and other desired statistical tests. Lastly, the data were sourced from two tertiary-care centers, which may have populations differing from the general populace in certain aspects (such as the prevalence of immunosuppression).

In conclusion, our findings regarding increased cavity wall thickness and the presence of perilesional consolidation in pulmonary lesions provide valuable insights into the diagnostic evaluation of these lesions. The association of increased cavity wall thickness with malignancy and the supportive role of perilesional consolidation in benignity underscore the importance of thorough radiological assessment in guiding clinical decision-making. Incorporating these imaging findings into the diagnostic algorithm can enhance the accuracy of diagnosing pulmonary lesions and facilitate appropriate patient management strategies.

## Data Availability

Data cannot be shared publicly because of Due to the law on the protection of personal rights. Data are owned by a third-party organization, (Research Ethics Committee of AIBU, contact email is boluetikibu.edu.tr).

---

**Table 1:** Receiver operating characteristic curves analysis maximum wall thickness associated with malignant cavities lesions

| Risc factor | AUC (95%) | Cutt of | P value | Sensitivity (%) | Spesivity (%) |
| --- | --- | --- | --- | --- | --- |
| Maximum wall thickness | 0.818<br>(0.714-0.921) | 23 | <0.001 | 44.7 | 97.1 |

## Notes

**Conflict of Interest:** None to declare.

### Competing Interest Statement

The authors have declared no competing interest.

### Funding Statement

The author(s) received no specific funding for this work.

### Author Declarations

This study was approved by Bolu Abant Izzet Baysal University ethics committee (2021-233)

